# Preliminary results of a randomized controlled trail: Effects of remotely supervised physical activity on health profile in frail older adults

**DOI:** 10.1101/2025.06.26.25330386

**Authors:** Xin Zhang, Feng Li, Jinwei Li, Xinyang Bai

## Abstract

**Introduction:** Supervised physical activity (PA) interventions have demonstrated efficacy and safety in improving health outcomes among frail populations. However, the growing disparity between limited healthcare resources and the rapidly aging population poses a significant challenge. Remotely delivered PA interventions may offer a viable solution for resource-limited regions. This study presents preliminary findings from a pre-designed protocol, examining the health benefits of a remotely supervised PA intervention, with a focus on frailty components.

**Methods:** Participants aged ≥65 years with a FRAIL score ≥3 were enrolled. The intervention group (IG) received a progressive PA program delivered via a WeChat-based application, along with smart insole-equipped sports shoes to monitor PA. The control group (CG) received routine care. The primary outcome was physical function, assessed via the Timed Up & Go (TUG) test; secondary outcomes included self-reported and objective measures. Assessments were conducted at baseline and 12 weeks post-intervention.

**Results:** Of the 20 participants enrolled (mean age 69 ± 5.73 years; 75% female), 3 withdrew. Baseline characteristics did not differ significantly between groups. Post-intervention, no significant between-group difference was observed in TUG (IG: 12.03 ± 0.39 vs CG:13.72 ± 0.85; *p* = 0.07). Generalized Estimating Equation (GEE) analysis confirmed a significant reduction in and FRAIL score (IG: 1.56 ± 0.23 vs CG: 2.88 ± 0.21; *p* < 0.001) for the IG.

**Discussion:** The 12-week remotely supervised PA intervention significantly improved frailty and physical performance but did not enhance psychological or cognitive function. Future large-scale studies are needed to comprehensively evaluate effectiveness, particularly long-term outcomes.

## 1. Introduction

Physical Activity (PA) plays a critical role in delaying and reversing the trajectory of frailty among older adults, and PA intervention has been widely conducted in the field of frailty management (1). It’s strongly recommended that a multi-component PA program, combining resistance with aerobic and balance training, should be offered and available for older adults with frailty (2). Emerging evidence further suggests that considering personalized preference and combining progressive goal-setting strategy might enhance long-term adherence (3, 4).

Supervised PA intervention has been proven to provide more additional effectiveness over unsupervised programs, particularly in the premise of ensuring the safety, while upholding the training intensity (5-7). Traditionally supervised PA programs for older adults are typically center-based, which means participants need to commute to training facilities on training days (8). However, this approach poses potential fall risks, particularly under adverse weather conditions, such as icy or snow covered roads in northern China (9). In such contexts, home-based interventions serve as a safer and more feasible alternative.

Nevertheless, the main challenge is the imbalance of limited healthcare providers and the rapidly growing population of older adults, especially in the regions with limited resource (10). Technology development has opened up new possibilities for home-based remotely supervised intervention among general community-dwelling older adults (11). Older adults also showed acceptable interest in platform-based intervention (12), and the initial pilot study has indicated promising feasibility and participation adherence (13). In China, WeChat is the most widely used multifunctional social platform across all age groups (14). Given its extensive reach, intervention delivered by WeChat is considered as a highly accessible and cost-effectiveness approach (15).

Despite the well-documented benefits of PA programs in improving frailty status and enhancing multiple functional aspects (16), the efficacy of remotely supervised PA interventions on frailty status remains unclear, particularly regarding their specific effects on individual frailty components. Establishing the effectiveness of technology-based remote PA supervision could facilitate its widespread implementation in large populations. Furthermore, understanding the progression of improvements across frailty components is crucial for investigating the interrelationships between physical and psychological domains. Such insights would provide valuable evidence for optimizing targeted intervention strategies in future implementations. Therefore, the primary purpose of this study is to compare the efficiency of on-site and remotely supervised PA programs. Secondly, to examine the impact of remotely supervised PA programs for each frailty component. We hypothesize that both interventions will achieve approximately the same effect, with particular emphasis on the functional walking component potentially showing more improvements, suggesting that remote supervision can achieve results that are at least as effective as on-site supervision.

## 2. Methods

*Participants* Aligning with the study protocol (17), eligible participants aged 65 years or older were enrolled upon meeting the criterion of a FRAIL score ≥3. Stratified randomization was initiated once any of the six community centers successfully recruited 20 qualifying participants. All participants provided the signed written informed consent. This study is the preliminary report of the results for the first full-recruited community center (**Figure 1**).

**Figure 1.**
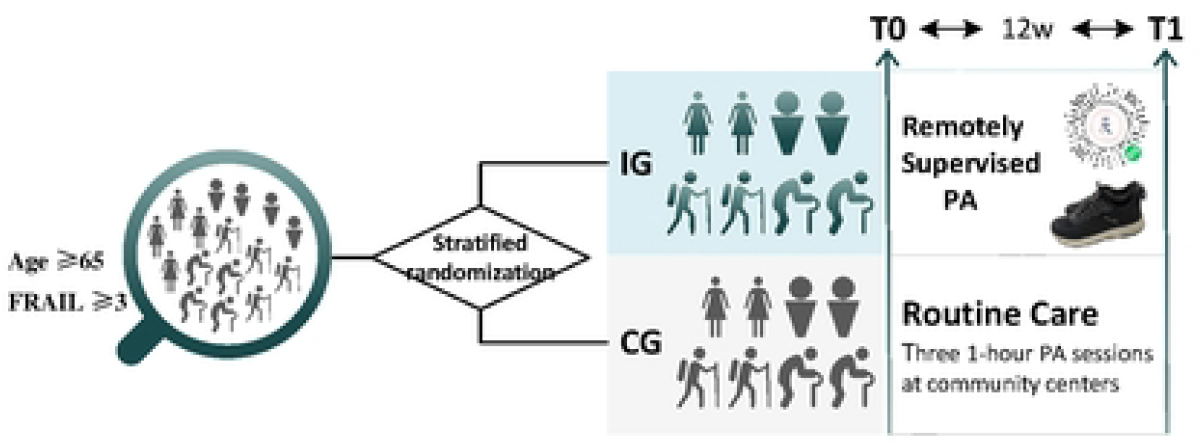
Flow Diagram.

*Intervention* A progressive PA program was designed for the intervention group (IG). The target PA goal was individualized based on each participant’s current PA level, with tailed training plan specifying the required frequency and intensity. Regarding the specific PA type and training day, it was decided by participants according to their own personal preference. The PA goal was adjusted weekly, with the initial goal-setting conducted during the first face-to-face interview assisted by a healthcare provider. The subsequent 11 goal adjustments were independently completed by participants via a WeChat-based application. Additionally, a pair of sport shoes equipped with the smart insoles inside was delivered to the participants at the first interview to record the PA. Participants were required to wear the shoes during PA sessions and, whenever possible, throughout the day except for sleeping, showering, and engaging in water-related activities. In the current study, the smart insole served only a monitoring purpose, the collected data were not utilized for analysis or participant feedback.

*Control* The Control Group (CG) received routine care during the intervention period, consisting of: three 1-hour PA sessions at community centers (including 3.6 km fast walking, aerobic dancing, and Pilates), and three health education lectures. These activities were scheduled every half-month on Wednesdays. These activities were routinely conducted by the community center, so we did not assess participant adherence in the CG.

*Outcomes* The primary outcome was physical function, measured by Time-Up & Go test (TUG), secondary outcomes covered both self-reported and objective measures. Due to practical considerations during study implementation, we modified the original protocol by using the FRAIL Scale instead of the planned Fried Phenotype to assess frailty status. Self-reported outcomes, collected through corresponding questionnaires, including weekly Moderate-to-Vigorous Physical Activity (MVPA), anxiety symptoms, and cognitive function. Objective measures comprised Short Physical Performance Battery (SPPB), gait speed evaluated by 4m walking test, and gait variability identified as the coefficient variation of stride time.

Gait variability was evaluated during a 2-minute walking test in a 10-meter pace. Participants were required to walk by their habitual walking speed, and completed the necessary walking condition, including go straight, turning, acceleration, and deceleration. The data was obtained using a medical system named “NOITOM”. Seven inertial measurement units (IMUs) were positioned by band at: the posterior lumbosacral area (midpoint of the intercrestal line), the lateral aspects of both thighs (the upper third line), the anterior aspects of both calves (Proximal popliteal skin crease with knee flexed at 90°), the dorsal surfaces of both feet (**Figure 2**). Details about other measures, please refer to the proposal (17).

**Figure 2.**
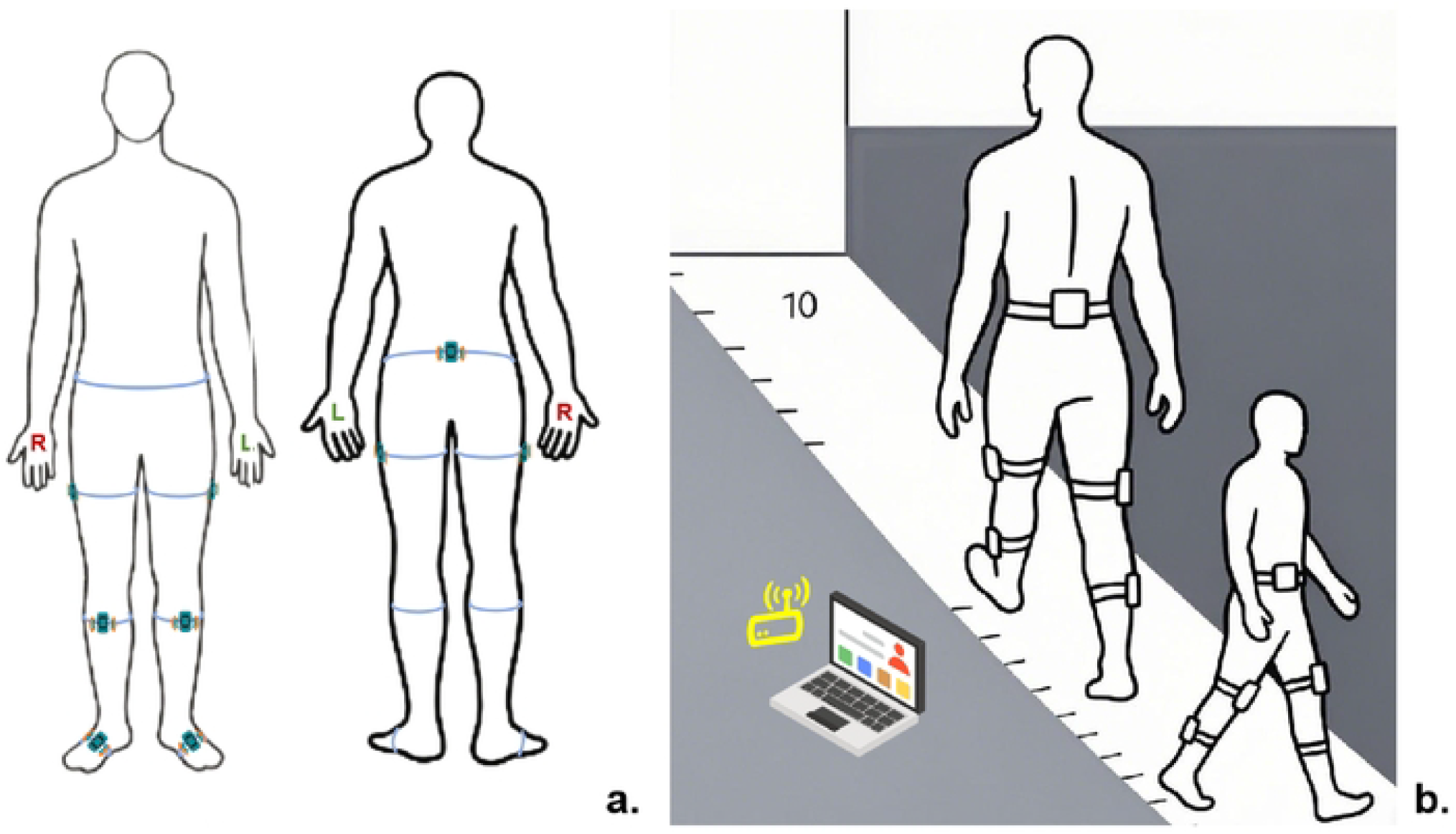
Illustration of “NOITOM” system. Panel a. Attached position of IMU; Panel b. 2-minutes walking test in a 10-meter pace.

All assessments were conducted at baseline and 12 weeks post-intervention. In accordance with ethical considerations, as the control group was scheduled to receive the intervention after study completion, follow-up data are not reported in this paper. Details about measurements, please refer to the proposal (17).

*Statistical Analysis* Normally distributed numerical variables were presented as mean ± standard deviation and analyzed using t-tests for between-group comparisons when met normality and homogeneity of variance assumptions; otherwise, the nonparametric Mann-Whitney U test was operated. Categorical variables were described using frequencies (percentages) and compared between groups using chi-square tests. To examine interaction effects between grouping factors and time factors, Generalized Estimating Equations (GEE) model were promoted.

## 3. Results

From May 2023, 495 individuals were screened, 43 participants meet the inclusion criteria. The first community center included 20 participants, and started the randomization and intervention, with 10 subjects assigned to each group: intervention group (IG) and control group 136 (CG).

### 3.1 General characteristics

The mean age of the participants was 69 years (SD: 5.73), most of them were female (75%), married and living with their partner (75%), finished 12-year senior-high school education (75%), and reported no fall history in the last 12 months (80%). Additionally, most of them were non-smokers and alcohol-free (90%), and took fewer than 3 drugs (75%). Regarding the components of frailty, all the participants with FRAIL score of 3 at baseline, the most prevalent component is fatigue (90%), followed by weight loss (85%), comorbidities (65%), functional resistance (40%), and functional walking (10%). No significant differences were observed across demographic characteristics and frailty components between IG and CG (**Table 1**).

**Table 1.**
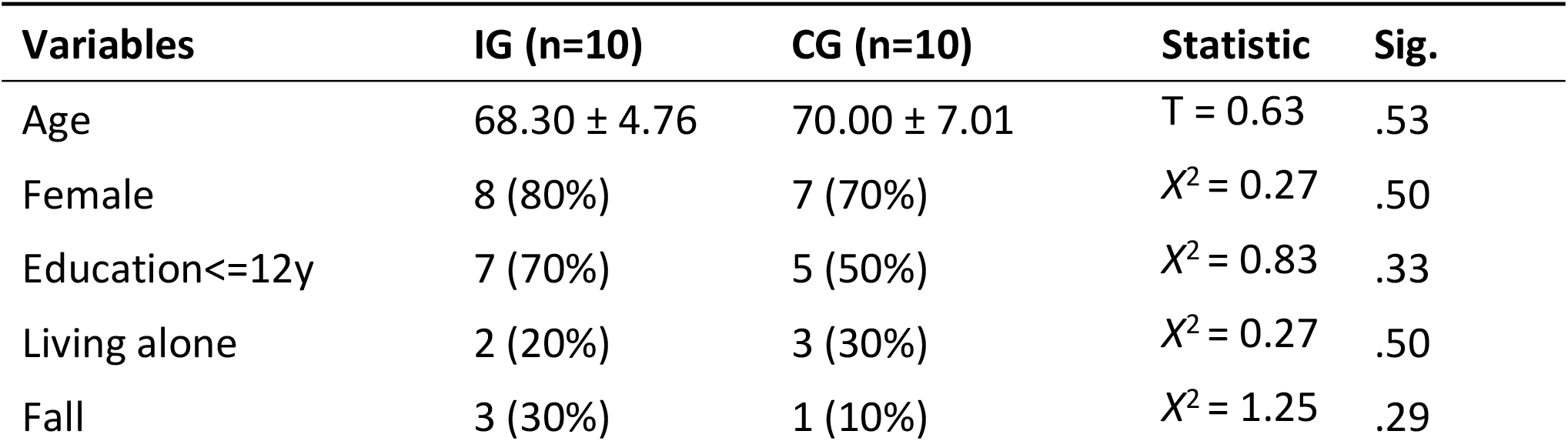
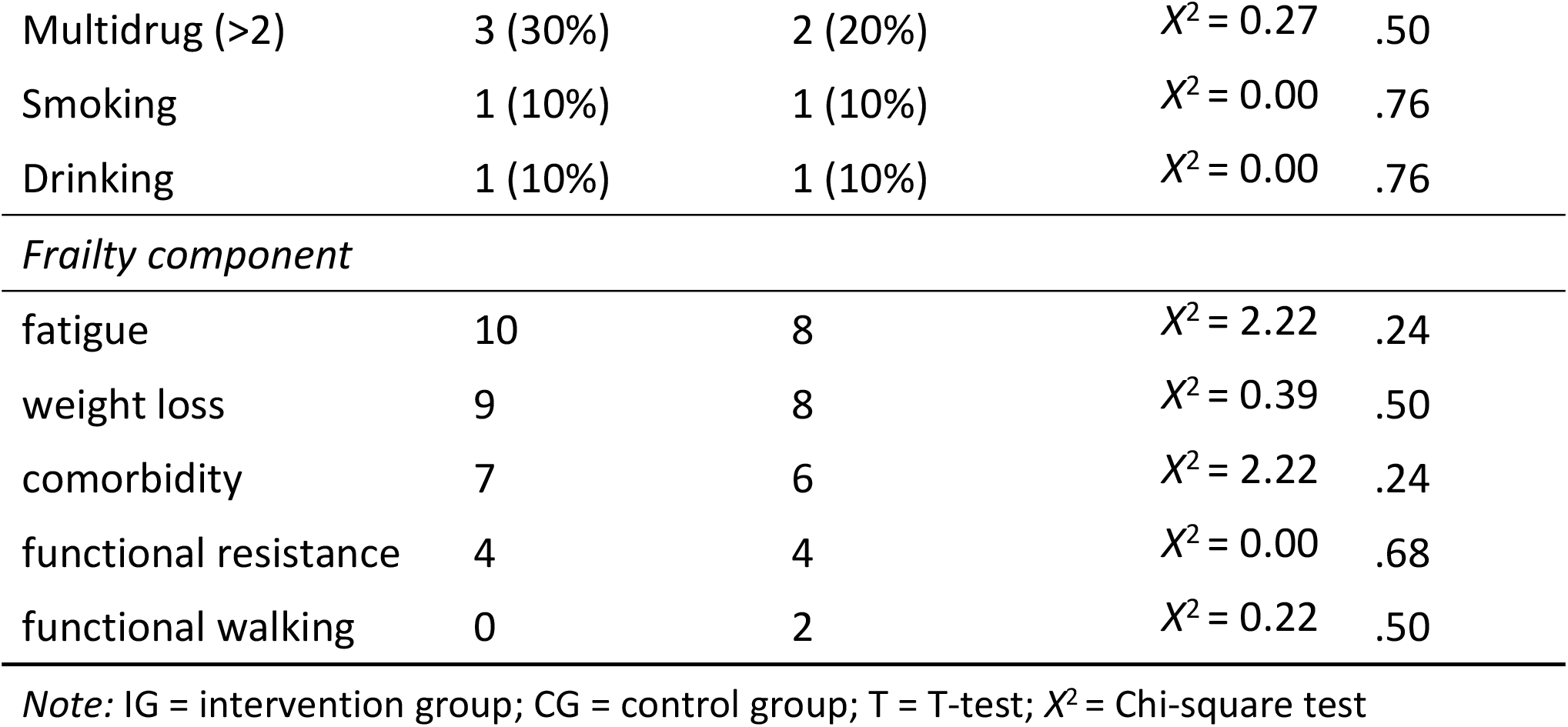
Comparation of demographic characteristics and frailty components at baseline.

### 3.2 Adherence to the study protocol

Between April to June in 2024, a total of 17 participants completed a 12-week intervention, with 3 withdrawals recorded (IG: n=1; CG: n=2).

### 3.3 Baseline and post-intervention difference between IG and CG

**Table 2** presents the mean values of the observed outcomes. No significant differences were observed between IG and CG at baseline. At post-intervention, significant differences were observed in FRAIL score, SPPB, and gait variability between IG and CG.

In IG, functional performance outcomes, including gait speed, Short Physical Performance Battery (SPPB) and Mini Mental State Examination (MMSE) increased at post-intervention, while gait variability and Generalized Anxiety Disorder-7 (GAD7) decreased. Conversely, the CG group showed opposing trends in these measures, except for MMSE.

**Table 2.**
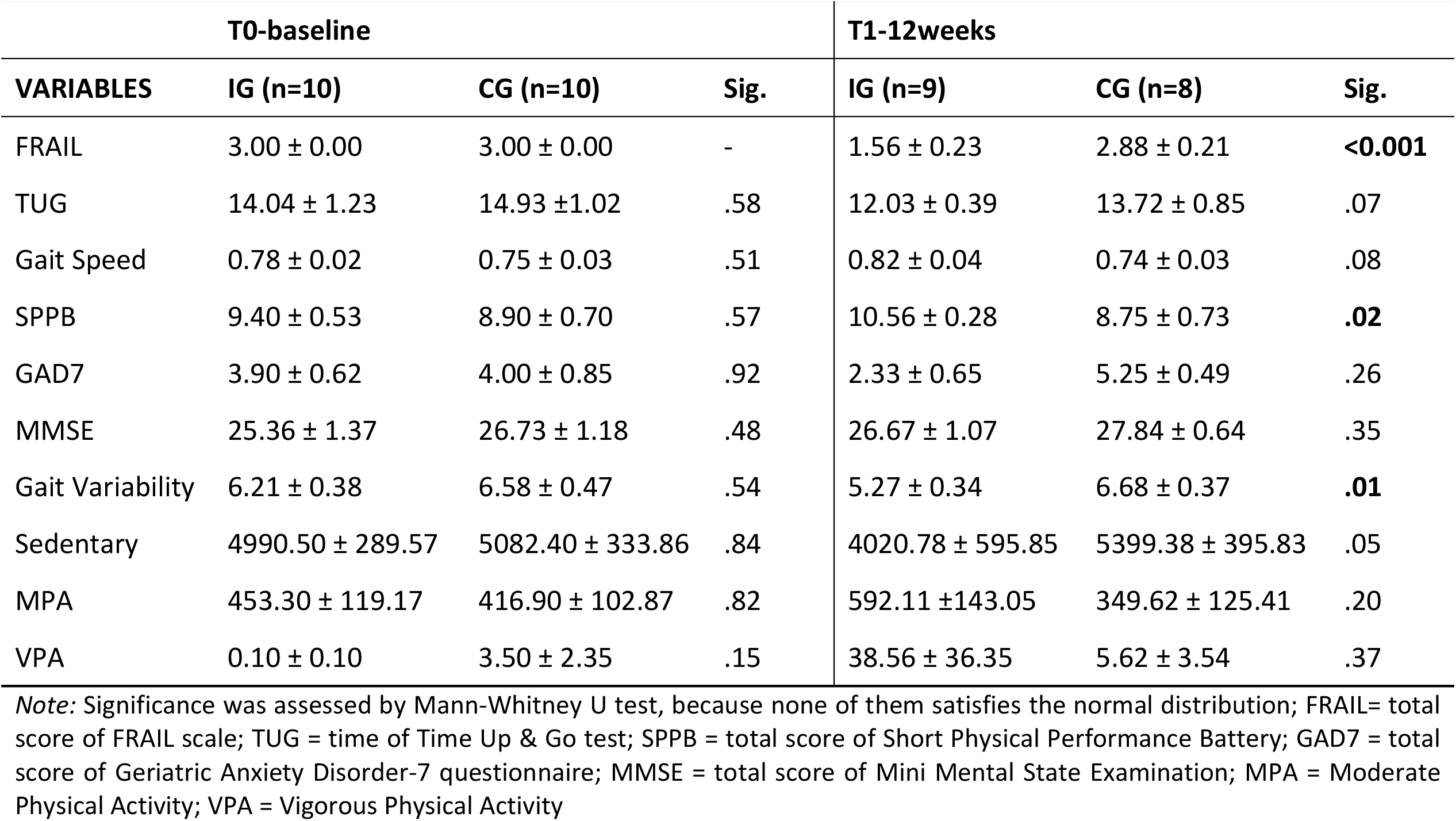
Pairwise Comparation between groups at T0 and T1.

Regarding PA behaviors, IG group showed increased Moderate-to-Vigorous Physical Activity (MVPA). While VPA also rose in CG group, but MPA declined. Sedentary activity decreased in the IG group but increase in CG group.

The results of Generalized Estimating Equation (GEE) analysis indicated the significant reduction in mean FRAIL scores, the performance of Time-Up & Go (TUG) test, the score of Short Physical Performance Battery (SPPB), and Geriatric Anxiety Disorder-7 (GAD7). The particularly significant reduction of FRAIL score was observed in IG group (*p*<0.001). Additionally, between-group differences were observed in gait variability (**Table 3**).

**Table 3.**
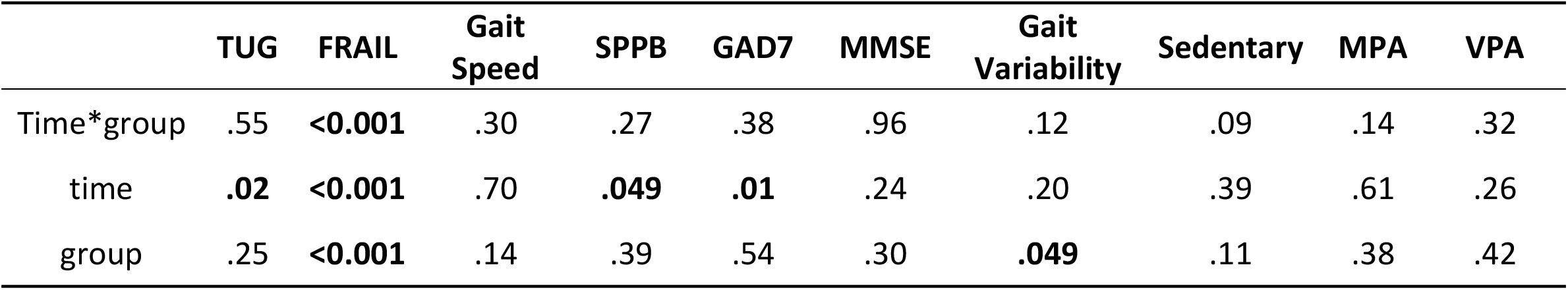
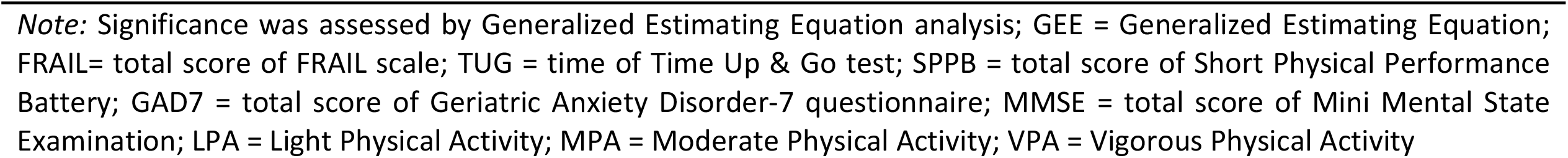
Comparation between and within groups (GEE)

|  | TUG | FRAIL | Gait Speed | SPPB | GAD7 | MMSE | Gait Variability | Sedentary | MPA | VPA |
| --- | --- | --- | --- | --- | --- | --- | --- | --- | --- | --- |
| Time*group | .55 | **<0.001** | .30 | .27 | .38 | .96 | .12 | .09 | .14 | .32 |
| time | **.02** | **<0.001** | .70 | **.049** | **.01** | .24 | .20 | .39 | .61 | .26 |
| group | .25 | **<0.001** | .14 | .39 | .54 | .30 | **.049** | .11 | .38 | .42 |

Note: Significance was assessed by Generalized Estimating Equation analysis; GEE = Generalized Estimating Equation; FRAIL= total score of FRAIL scale; TUG = time of Time Up & Go test; SPPB = total score of Short Physical Performance Battery; GAD7 = total score of Geriatric Anxiety Disorder-7 questionnaire; MMSE = total score of Mini Mental State Examination; LPA = Light Physical Activity; MPA = Moderate Physical Activity; VPA = Vigorous Physical Activity

### 3.4 Changes in the frailty components

After the intervention, improvement in FRAIL trajectory were observed in eight participants from IG, while only two improvements but one deterioration exhibited in CG group. Among the ten cases showing improvement, reductions in self-reported fatigue (n=7), functional resistance (n=4), weight loss (n=3), functional walking capacity (n=1) and comorbidities (n=1). The single case of deterioration in the CG was linked to new-onset self-reported exhaustion. Interestingly, the sole participant exhibiting improved functional walking capacity (in the CG) also reported persistent exhaustion (***Figure 3***).

**Figure 3.**
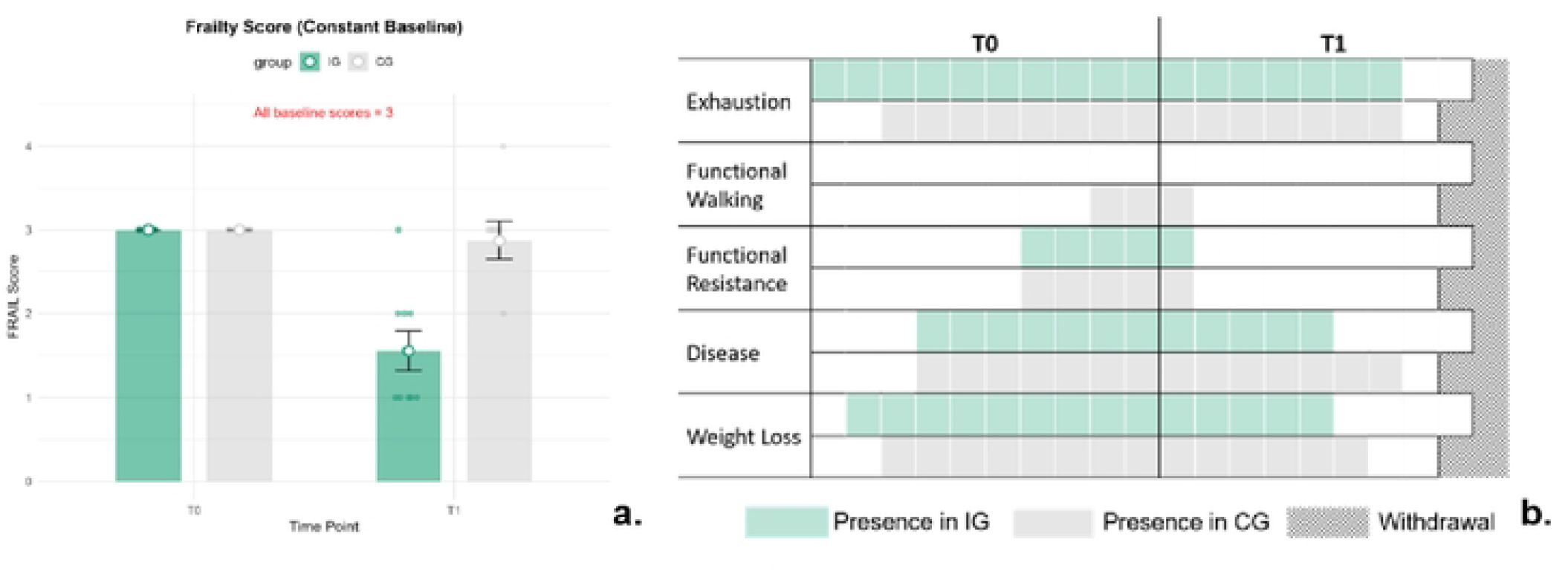
Results of FRAIL score and each component. Panel a. Bar plot of FRAIL score between IG and CG group in T0 and T1, dot represents the specific FRAIL score of each case; Panel b. Clustered column chart of the presence of five frailty components at T0 and T1, one square represents one subject.

### 3.3 Secondary outcomes

Among the nine participants in the intervention group (IG) who completed the full program, the averaged pass rate of the targeted goal is 7, with five (55.6%) sustained the target goal for over eight weeks.

## 4. Discussion

The preliminary result suggest that 12-week remotely supervised PA intervention achieved the comparable effectiveness for the health profile among frail older adults, significantly improved the frailty syndrome, especially the fatigue, as well as physical performance, but no significant for psychological and cognitive function.

While significant time effects were observed for both physical function (assessed via TUG and SPPB) and psychological function (measured by anxiety symptoms), no significant time × group interaction effects were detected. This lack of interaction may be attributable to the limited sample size at the current stage of research. Nevertheless, a positive improvement trajectory was noted in the IG, suggesting preliminary efficacy of the intervention, which has been demonstrated in previous studies (18, 19). Notably, improvements in TUG performance were less pronounced compared to SPPB scores, potentially due to the inherently smaller magnitude of change associated with TUG measures (20). This observation underscores the need for careful interpretation of functional outcomes when using different assessment tools.

The IG demonstrated increases in both MPA and VPA, accompanied by a reduction in sedentary behavior. In contrast, the CG showed an increase only in VPA, with decreases observed in the other two activity categories. Due to the inherent limitations of the IPAQ measurement tool, changes in light physical activity (LPA) could not be directly assessed. However, we hypothesize that LPA likely increased in the IG based on the principle of 24-hour activity redistribution (13, 21). Regarding the CG, the observed VPA increase may be attributed to the scheduled 1-hour PA sessions at community centers, particularly as post-intervention assessments were conducted immediately following these sessions. This temporal proximity suggests potential acute effects rather than sustained behavioral changes.

Align with the previous research findings (13), PA programs can mitigate frailty by targeting physical endurance and perceived exhaustion (22). The frail level was improved after the intervention, reflected by the decline of FRAIL score (23), who attributed similar effects to enhanced neuromuscular adaptation and energy metabolism. The improvement observed above suggests that perceived well-being may be the first aspect to demonstrate positive changes, subsequently influencing a broader health profile. This finding contrasts with our initial hypothesis that functional walking capacity would show the most significant gains, despite most intervention programs incorporating targeted lower extremity training.

However, the bidirectional relationship between fatigue and physical activity levels highlights an important consideration: PA intervention programs should account for participants’ baseline physical status. Without proper individualization, high-intensity physical activity may inadvertently lead to fatigue and subsequent compensatory behaviors, such as prolonged sedentary periods, potentially offsetting the intended benefits (24). The observed negative trend in the CG may be partially attributable to unmonitored adherence to routine care. This finding underscores the critical advantage of remotely supervised PA interventions, which enable program fidelity.

The remotely delivered physical activity program demonstrated acceptable feasibility based on satisfactory adherence rates and goal achievement. However, the current small sample size increases Type II error risk and may mask potential intervention effects (25), and a more comprehensive analysis will be conducted upon study completion. The limited improvements in cognitive function may suggest the need for extended intervention periods, particularly for addressing cognitive frailty (26). While psychological improvements may benefit from social support mechanisms, incorporating group-based elements may provide additional effects (27). For long-term sustainability, integrating behavioral nudges (e.g., gamification) could help address the engagement challenges characteristic of remote programs (28).

## 6. Conclusion

The preliminary findings provide initial evidence supporting the feasibility and acceptability of the remotely delivered physical activity intervention. A comprehensive evaluation of its effectiveness, particularly the long-term efficiency, requires completion of full study.

## Data Availability

Data cannot be shared publicly because of ethic consideration. Data are available from the Ethics Committee for researchers who meet the criteria for access to confidential data.

## 7. Declaration

### Ethics approval and content of participate

This study was approved by the Human Research Ethics Committee of the School of Nursing, Jilin University (HREC 2020122001). All participants will provide a signed informed consent when entering the study.

### Availability of data and materials

The dataset supporting the conclusion of this article is available in the corresponding authors.

### Author contributions

Conceptualization and methodology: Xin Zhang, Jinwei Li, Li Feng Li. Data curation: Xin Zhang, Feng Li.

Formal analysis and interpretation of data: all authors.

Drafting of the manuscript (writing and visualization): Xin Zhang, Feng Li

Critical revision of the manuscript for important intellectual content: all authors.

## Acknowledgements

We acknowledge all members in team “Alpha Care” for their contribution to the measurement.

## References

1. Marcucci M, Damanti S, Germini F, Apostolo J, Bobrowicz-Campos E, Gwyther H, et al. Interventions to prevent, delay or reverse frailty in older people: a journey towards clinical guidelines. BMC Med. 2019;17(1):193.

2. Dent E, Morley JE, Cruz-Jentoft AJ, Woodhouse L, Rodríguez-Mañas L, Fried LP, et al. Physical Frailty: ICFSR International Clinical Practice Guidelines for Identification and Management. J Nutr Health Aging. 2019;23(9):771–87.

3. Barrado-Martín Y, Frost R, Catchpole J, Rookes T, Gibson S, Avgerinou C, et al. Goal setting as part of a holistic intervention to promote independence in older people with mild frailty: a process evaluation alongside a randomised controlled trial. Lancet. 2023;402 Suppl 1:S1.

4. Noone J, Mucinski JM, DeLany JP, Sparks LM, Goodpaster BH. Understanding the variation in exercise responses to guide personalized physical activity prescriptions. Cell Metab. 2024;36(4):702–24.

5. Karmakar P, Wong MC, AlMarzooqi MA, Alghamdi N, Ou K, Duan Y, et al. Enhancing Physical and Psychosocial Health of Older Adults in Saudi Arabia through Walking: Comparison between Supervised Group-Based and Non-Supervised Individual-Based Walking. Eur J Investig Health Psychol Educ. 2023;13(11):2342–57.

6. Suikkanen S, Soukkio P, Kautiainen H, Kääriä S, Hupli MT, Sipilä S, et al. Changes in the Severity of Frailty Among Older Adults After 12 Months of Supervised Home-Based Physical Exercise: A Randomized Clinical Trial. J Am Med Dir Assoc. 2022;23(10):1717.e9-.e15.

7. Gómez-Redondo P, Valenzuela PL, Morales JS, Ara I, Mañas A. Supervised Versus Unsupervised Exercise for the Improvement of Physical Function and Well-Being Outcomes in Older Adults: A Systematic Review and Meta-analysis of Randomized Controlled Trials. Sports Med. 2024;54(7):1877–906.

8. Chen YM, Berkowitz B. Older adults’ home- and community-based care service use and residential transitions: a longitudinal study. BMC Geriatr. 2012;12:44.

9. Yang M, Gao Q, Li T. Causes of Winter Persistent Extreme Cold Events in Northeastern China. Advances in Atmospheric Sciences. 2025;42(4):780–93.

10. Ashley L, Boussebaa M, Friedman S, Harrington B, Heusinkveld S, Gustafsson S, et al. Professions and inequality: Challenges, controversies, and opportunities. Journal of Professions and Organization. 2022;10(1):80–98.

11. Timm L, Guidetti S, Taloyan M. POSITIVE: experiences of an intervention aiming for reversing and preventing frailty using a home monitoring and communication platform within primary health care. BMC Geriatr. 2024;24(1):382.

12. Li N, Liu C, Wang N, Lin S, Yuan Y, Huang F, et al. Feasibility, usability and acceptability of a lifestyle-integrated multicomponent exercise delivered via a mobile health platform in community-dwelling pre-frail older adults: a short-term, mixed-methods, prospective pilot study. BMC Geriatr. 2024;24(1):926.

13. Li N, Wang N, Xu Y, Lin S, Yuan Y, Huang F, et al. The impacts of a mHealth platform-enabled lifestyle-integrated multicomponent exercise program on reversing pre-frailty in community-dwelling older adults: A randomized controlled trial. Int J Nurs Stud. 2025;167:105072.

14. Wei H. WeChat now an integral part of daily life in China, says survey China Daily 2021 [

15. Wang J, Zeng Z, Dong R, Sheng J, Lai Y, Yu J, et al. Efficacy of a WeChat-based intervention for adherence to secondary prevention therapies in patients undergoing coronary artery bypass graft in China: A randomized controlled trial. J Telemed Telecare. 2022;28(9):653–61.

16. Teh R, Barnett D, Edlin R, Kerse N, Waters DL, Hale L, et al. Effectiveness of a complex intervention of group-based nutrition and physical activity to prevent frailty in pre-frail older adults (SUPER): a randomised controlled trial. Lancet Healthy Longev. 2022;3(8):e519–e30.

17. Zhang X, Li J, Sui X, Xu L, Zhu L, Pang Y, et al. Effects of Remotely Supervised Physical Activity on Health Profile in Frail Older Adults: A Randomized Controlled Trial Protocol. Front Aging Neurosci. 2022;14:807082.

18. Zika MA, Becker L. Physical Activity as a Treatment for Social Anxiety in Clinical and Non-clinical Populations: A Systematic Review and Three Meta-Analyses for Different Study Designs. Frontiers in Human Neuroscience. 2021;Volume 15-2021.

19. Weng W-H, Cheng Y-H, Yang T-H, Lee S-J, Yang Y-R, Wang R-Y. Effects of strength exercises combined with other training on physical performance in frail older adults: A systematic review and meta-analysis. Archives of Gerontology and Geriatrics. 2022;102:104757.

20. Friedrich B, Lau S, Elgert L, Bauer JM, Hein A. A Deep Learning Approach for TUG and SPPB Score Prediction of (Pre-) Frail Older Adults on Real-Life IMU Data. Healthcare. 2021;9(2):149.

21. Wanigatunga AA, Cai Y, Urbanek JK, Mitchell CM, Roth DL, Miller ER, et al. Objectively Measured Patterns of Daily Physical Activity and Phenotypic Frailty. J Gerontol A Biol Sci Med Sci. 2022;77(9):1882–9.

22. Cadore EL, Casas-Herrero A, Zambom-Ferraresi F, Idoate F, Millor N, Gómez M, et al. Multicomponent exercises including muscle power training enhance muscle mass, power output, and functional outcomes in institutionalized frail nonagenarians. Age (Dordr). 2014;36(2):773–85.

23. Nagai K, Miyamato T, Okamae A, Tamaki A, Fujioka H, Wada Y, et al. Physical activity combined with resistance training reduces symptoms of frailty in older adults: A randomized controlled trial. Arch Gerontol Geriatr. 2018;76:41–7.

24. Fanning J, Nicklas BJ, Rejeski WJ. Intervening on physical activity and sedentary behavior in older adults. Experimental Gerontology. 2022;157:111634.

25. Button KS, Ioannidis JPA, Mokrysz C, Nosek BA, Flint J, Robinson ESJ, et al. Power failure: why small sample size undermines the reliability of neuroscience. Nature Reviews Neuroscience. 2013;14(5):365–76.

26. Yuan Y, Wang S, Zhou C, Zhang A, Zhang S, Wang Y. Effects of exercise interventions on cognition, physical function and quality of life among older adults with cognitive frailty: A systematic review and meta-analysis. Geriatric Nursing. 2025;62:96–107.

27. Gen A, Higuchi Y, Ueda T, Hashimoto T, Kozuki W, Murakami T, et al. Intervention for Social Frailty Focusing on Physical Activity and Reducing Loneliness: A Randomized Controlled Trial. Clin Interv Aging. 2025;20:43–53.

28. Avgerinou C, Gardner B, Kharicha K, Frost R, Liljas A, Elaswarapu R, et al. Health promotion for mild frailty based on behaviour change: Perceptions of older people and service providers. Health Soc Care Community. 2019;27(5):1333–43.

